# Decoding hand and wrist movement intention from chronic stroke survivors with hemiparesis using a user-friendly, wearable EMG-based neural interface

**DOI:** 10.1101/2021.09.07.21262896

**Authors:** Eric C Meyers, David Gabrieli, Nick Tacca, Lauren Wengerd, Michael Darrow, David Friedenberg

**Author notes:** These authors contributed equally (Eric C Meyers, David Gabrieli).

## Abstract

Prosthetics and orthotics have been recognized for decades as a potential means to restore hand function and independence to individuals living with impairment due to stroke. However, 75% of stroke survivors, caregivers, and health care professionals (HCP) believe current practices are insufficient, specifically calling out the upper extremity as an area where innovation is needed to develop highly usable prosthetics/orthotics for the stroke population. A promising method for controlling upper limb technologies is to infer movement intent non-invasively from surface electromyography (EMG) activity. While this approach has garnered significant attention in the literature, existing technologies are often limited to research settings and struggle to meet stated user needs. To address these limitations, we have developed the NeuroLife^®^ EMG System, which consists of a wearable forearm sleeve with 150 embedded electrodes and associated hardware and software to record and decode surface EMG. Here, we demonstrate accurate decoding of 12 functional hand, wrist, and forearm movements, including multiple types of grasps from participants with varying levels of chronic impairment from stroke, with an overall accuracy of 77.1±5.6%. Importantly, we demonstrate the ability to decode a subset of 3 fundamental movements in individuals with severe hand impairment at 85.4±6.4% accuracy, highlighting the potential as a control mechanism for assistive technologies. Feedback from stroke survivors who tested the system indicates that the sleeve’s design meets various user needs, including being comfortable, portable, and lightweight. The sleeve is in a form factor such that it can be used at home without an expert technician and can be worn for multiple hours without discomfort. Taken together, the NeuroLife EMG System represents a platform technology to record and decode high-definition EMG for the eventual real-time control of assistive devices in a form factor designed to meet user needs.

## Introduction

Stroke is a leading cause of long-term disability in the United States, affecting more than 800,000 people per year [1]. Unilateral paralysis (hemiparesis) affects up to 80% of stroke survivors, leaving many to struggle with activities of daily living (ADLs) involving manipulating objects such as doors, utensils, and clothing due to decreased upper-limb muscle coordination and weakness [2]. Restoration of hand and arm function to improve independence and overall quality of life is a top priority for stroke survivors and caregivers [3]. Intensive physical rehabilitation is the current gold standard for improving motor function after stroke. Unfortunately, 75% of stroke survivors, caregivers, and health care providers report that current upper extremity training practice is insufficient [4]. The development of user-centric neurotechnologies to restore motor function in stroke survivors could address these unmet clinical needs through a range of different mechanisms such as improving motivation, enhancing neuroplasticity in damaged sensorimotor networks, and enabling at-home therapy.

Assistive technologies (AT) hold potential to restore hand function and independence to individuals with paralysis [5]. ATs, including exoskeletons and functional electrical stimulation (FES), can assist with opening the hand and also evoke grips strong enough to hold and manipulate objects [6]. Additionally, these systems have been used therapeutically during rehabilitation to strengthen damaged neural connections to restore function [7]. A wide variety of mechanisms to control ATs have been investigated including voice [8], switch [9], position sensors [10], electroencephalography (EEG) [11], electrocorticography (ECoG) [12], intracortical microelectrode arrays (MEA) [13], and electromyography (EMG) [14]. Unfortunately, no single system has simultaneously delivered an intuitive, user-friendly system with a high degree-of-freedom (DoF) control for practical use in real-world settings [4].

Recent advances in portable, high-definition EMG-based (HDEMG) systems have the potential to overcome several of these barriers and deliver an intuitive and entirely non-invasive AT control solution [15,16]. While various EMG-based ATs exist, including the MyoPro Orthosis that is commercially available [15], most of these systems use a small number of electrodes and rely on threshold-based triggering [14]. Consequently, these systems have limited DoF control which limits their practical use. Conversely, HDEMG systems leveraging machine learning approaches to infer complex movement intention can provide high DoF control, significantly expanding functional use cases and the proportion of the stroke population that could benefit from these technologies [16,17]. Currently, HDEMG systems are primarily confined to the research setting and have usability limitations, including being difficult to set up, requiring manual placement of electrodes, and being non-portable and bulky, which can hinder the successful translation of technologies [4].

To address these limitations, we have recently developed the NeuroLife^®^ EMG System to decode complex forearm motor intention in chronic stroke survivors while simultaneously addressing end user needs. The EMG system was designed to be used as a control device for various end effectors such as FES systems and exoskeletons. Additionally, the system was designed to meet user needs in the following domains that have previously been identified as high-value for stroke survivors: donning/doffing simplicity, device setup and initialization, portability, robustness, comfortability, size and weight, and intuitive usage [4]. The sleeve is a wearable garment consisting of up to 150 embedded electrodes that measure muscle activity in the forearm to decode the user’s motor intention. A single zipper on one edge of the sleeve allows for a simplified and streamlined donning and doffing by the user and/or a caregiver. The sleeve design facilitates an intuitive setup process as embedded electrodes are consistently placed on the arm eliminating the need for manual electrode placement on specific muscles. The lightweight stretchable fabric, similar to a compression sleeve, was chosen to enhance comfort for long-term use. Overall, these design features of the sleeve help address critical usability factors for ATs [4].

In this work, we demonstrate that our wearable EMG system can extract task-specific myoelectric activity at high temporal and spatial resolution to resolve individual movements. Based on EMG data collected from seven individuals with upper extremity hemiparesis due to stroke, trained neural network machine learning models can accurately decode muscle activity in the forearm to infer movement intention, even in the absence of overt motion. We assess the viability of this technique for real time decoding, demonstrating the feasibility of controlling ATs based on motor intention. Finally, we present usability data collected from the study participants that highlight the user-centric design of the sleeve. This data will be used to inform future developments to ultimately deliver an effective EMG-based neural interface that meets end user needs. Together, we present a capable, user-centric EMG-based neural interface for the detection of motor intention after stroke, with usability guidelines that will inform future device improvements.

## Methods

### Participants

Eight individuals (3 female, 4 male; 60±5 years) with a history of stroke participated in a study that recorded EMG using the NeuroLife EMG System while attempting various hand and wrist movements. Additionally, data was collected from seven able-bodied individuals (4 female, 3 male; 27±1 years) to serve as a general comparison of EMG data, and to benchmark decoding algorithms. Data was collected as part of an ongoing clinical study being conducted at Battelle Memorial Institute and approved by the Battelle Memorial Institute Institutional Review Board (IRB No. 0779 and IRB No. 0773, respectively). Informed consent was obtained from all participants prior to any experimental procedures. Demographics on study participants with stroke are provided in Table 1, and for able-bodied participants in Supplementary Table 1. Eligibility criteria were set to recruit adult chronic stroke survivors with hemiparesis affecting the arm and hand and who were able to follow 3-step commands and communicate verbally. Specific inclusion and exclusion criteria are listed in the Supplementary Methods.

**Table 1.** Demographics of participants with stroke.

| Participant | UEFM | UEFM-HS | Time since stroke, years | Side of paresis |
| --- | --- | --- | --- | --- |
| 13762 | 36 | 6 | 6 | Right |
| 29562 | 22 | 2 | 4 | Right |
| 30458 | 32 | 6 | 3 | Left |
| 47513 | 19 | 4 | 4 | Right |
| 61204 | 8 | 0 | 6 | Right |
| 87134 | 7 | 0 | 7 | Right |
| 98473 | 38 | 7 | 6 | Right |

During the first session prior to EMG data collection, standardized clinical assessments were performed by a licensed occupational therapist in all participants with stroke. These included the upper extremity section of the Fugl-Meyer (UE-FM) to assess upper extremity motor impairment, the Box and Blocks test to assess manual dexterity, and the Modified Ashworth test to assess spasticity of the finger, wrist, and elbow flexors. Based on predetermined exclusion criteria, one participant was removed from data analysis due to hemispatial neglect affecting their ability to consistently follow movement cues.

### Experimental Setup

Participants were seated facing a computer monitor with their arms placed on a table, and the sleeve was placed on the paretic arm for participants with stroke, and the right arm for able-bodied participants, regardless of handedness (Figure 1). The sleeve comprises a stretchable fabric with an embedded array of electrodes (Supplementary Figure 1). Depending on the forearm size of the participant, a small, medium, or large sized sleeve was used containing 128 electrodes (64 channel pairs), 142 electrodes (71 channel pairs), or 150 electrodes (75 channel pairs) respectively. Each electrode is 12mm diameter, spaced 25mm apart, and wrap the forearm from elbow to wrist. With a flexible and lightweight nylon-Lycra hybrid material, the sleeve wears like a compression sleeve and weighs 180, 195, and 220 grams for the small, medium, and large sleeves, respectively. A zipper on the ulnar edge of the sleeve allows for easy donning and doffing. Prior to donning, an electrode solution spray (Signaspray, Parker Laboratories, Fairfield, NJ) was applied to the participant’s forearm to improve signal quality. Bipolar EMG signals were sampled at 3KHz using an Intan Recording Controller (Intan Technologies, Los Angeles, CA). An embedded electrode in the sleeve near the elbow was used as a reference for all bipolar amplifiers. The sleeve was connected to a custom-built EMG signal acquisition module, which then connected to a laptop computer (Figure 1 and Supplementary Figure 1a).

**Figure 1.**
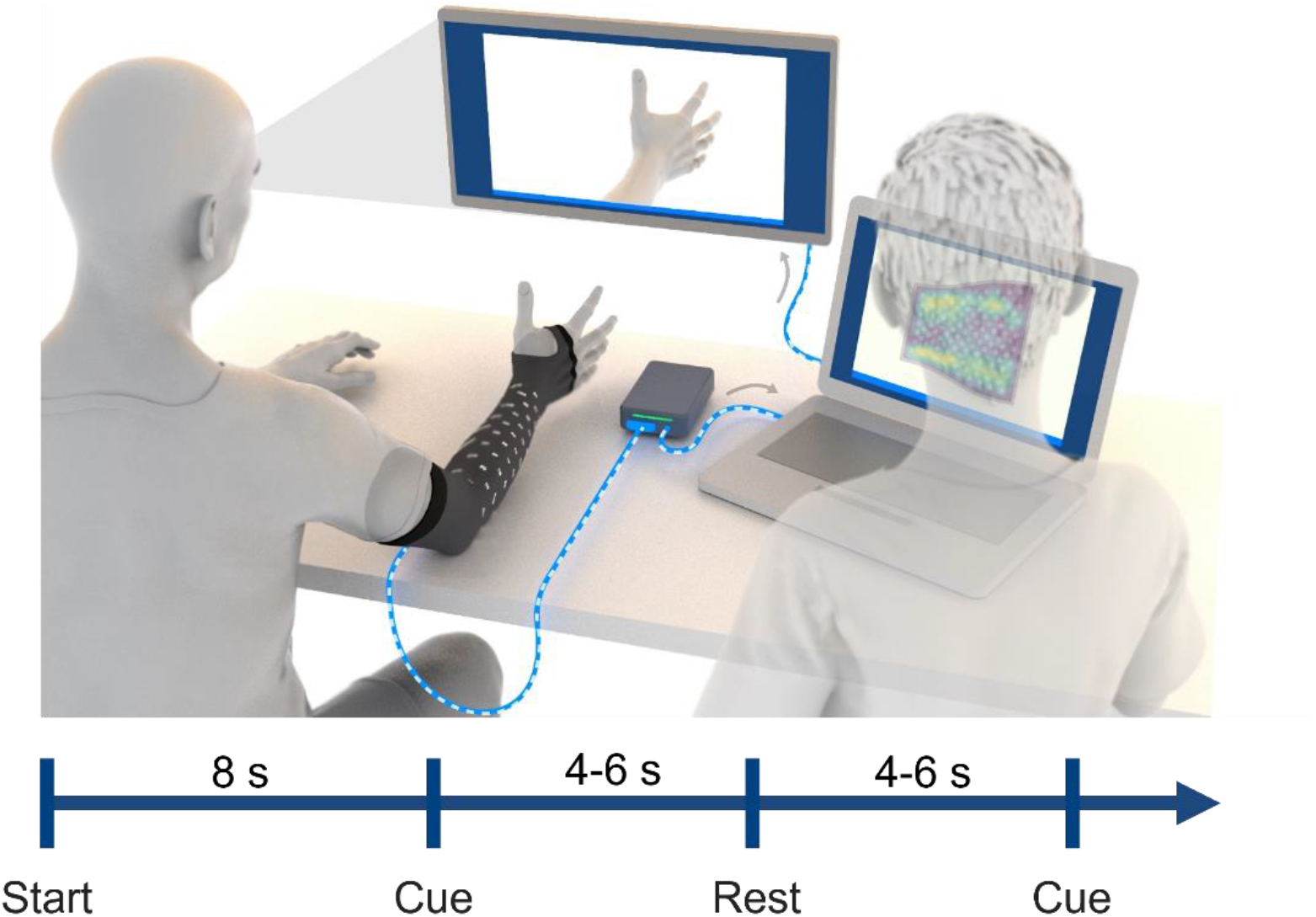
Illustration of experimental data collection procedure. Participants were seated in front of a computer monitor with the sleeve on their impaired arm, and their arms placed on the table. The sleeve was connected to a custom-built EMG signal acquisition module, which then connected to a laptop computer. Images of hand postures were shown on the monitor and the participant followed along to the best of their ability. Each recording block was approximately 2-3 minutes in length, and involved hand posture cues interleaved with rest periods. The recording block began with an 8-second lead in rest period. Each cue and rest period presentation time were randomly selected between 4-6 seconds for participants with stroke. An operator ran the data collection software and observed EMG signals during data collection to ensure proper recording of data.

The participants were instructed to attempt a series of hand, wrist, and forearm movements. A series of images of the desired hand movement was presented on a computer monitor, and the participants were instructed to attempt each movement shown to the best of their ability. Participants were instructed to attempt the movement at 25-50% maximal effort to minimize muscle fatigue and co-contractions throughout the session.

The following movements were collected during the session: Hand Flexion, Hand Extension, Index Extension, Thumb Flexion, Thumb Extension, Thumb Abduction, Forearm Supination, Forearm Pronation, Wrist Flexion, Wrist Extension, Two Point Pinch, and Key Pinch. These movements were identified by a licensed occupational therapist as highly relevant functional movements for dexterous hand use, and these movements have been used in similar studies [18]. Recording blocks consisted of a single movement repeated 10 times (referred to as “single blocks”), or multiple movements repeated within a single recording block (referred to as “mixed blocks”). Every block began with an 8s rest period, followed by alternating movement and rest periods. During mixed blocks, a collection of movements (e.g., hand flexion, hand extension, forearm supination) were randomly presented to the participant with interleaved rest periods. Before beginning the block, participants were shown the movement(s) in the upcoming block. For participants with stroke, the time for each movement was randomly selected from a uniform distribution between 4-6s, and rest time was randomly selected between 4-6s. For able-bodied participants, the movement and rest times were both set randomly between 2-3s. The cue and rest times were shortened in able-bodied participants due to faster movement times and the expectation of simpler decoding compared to the participants with stroke. In the last recording session, a usability questionnaire assessing user needs (adapted from [4]) was given to subjects to evaluate the usability of the current sleeve design (responses from participants are presented in Supplementary Table 3).

Data was collected across 3-4 sessions with each participant with stroke, and in 1 session with able-bodied participants. Each session was <2 hours. For participants with stroke, data from all sessions except the last half of the final session were used to train the classifiers. Total amount of training data per movement for participants with stroke are shown in Supplementary Figure 3. For able-bodied experiments, data was collected in a single session with a total of 10 repetitions for each movement with the first 5 repetitions used for training, and the last 5 repetitions used for testing, corresponding to 7.5 seconds of data used for both training and testing.

To assess each participant’s ability to perform the movements without any assistance, each movement was scored by a licensed occupational therapist based on a scoring scheme adapted from the Action Research Arm Test (ARAT) [18]. The “observed movement score” was ranked using the following categories: 0=no movement; 1=incomplete range of motion; 2=complete range of motion but impaired; 3=normal.

### Pre-processing, windowing, and feature extraction

The EMG data was bandpass filtered between 20-400Hz using a 10^th^ order Butterworth filter, and a 60Hz notch filter was applied similar to previous studies [20]. Following pre-processing, the root mean square (RMS) was extracted using 100ms data windows with no overlap. An example of the RMS extraction for multiple movement sets are shown in Figure 2B,C. For decoding of movement intent during a given time window, the current window and three preceding windows were used, totaling 400ms of RMS data used for each prediction. Next, the training data was normalized (mean=0, variance=1) and the testing data was normalized using the mean and variance from the training data.

**Figure 2.**
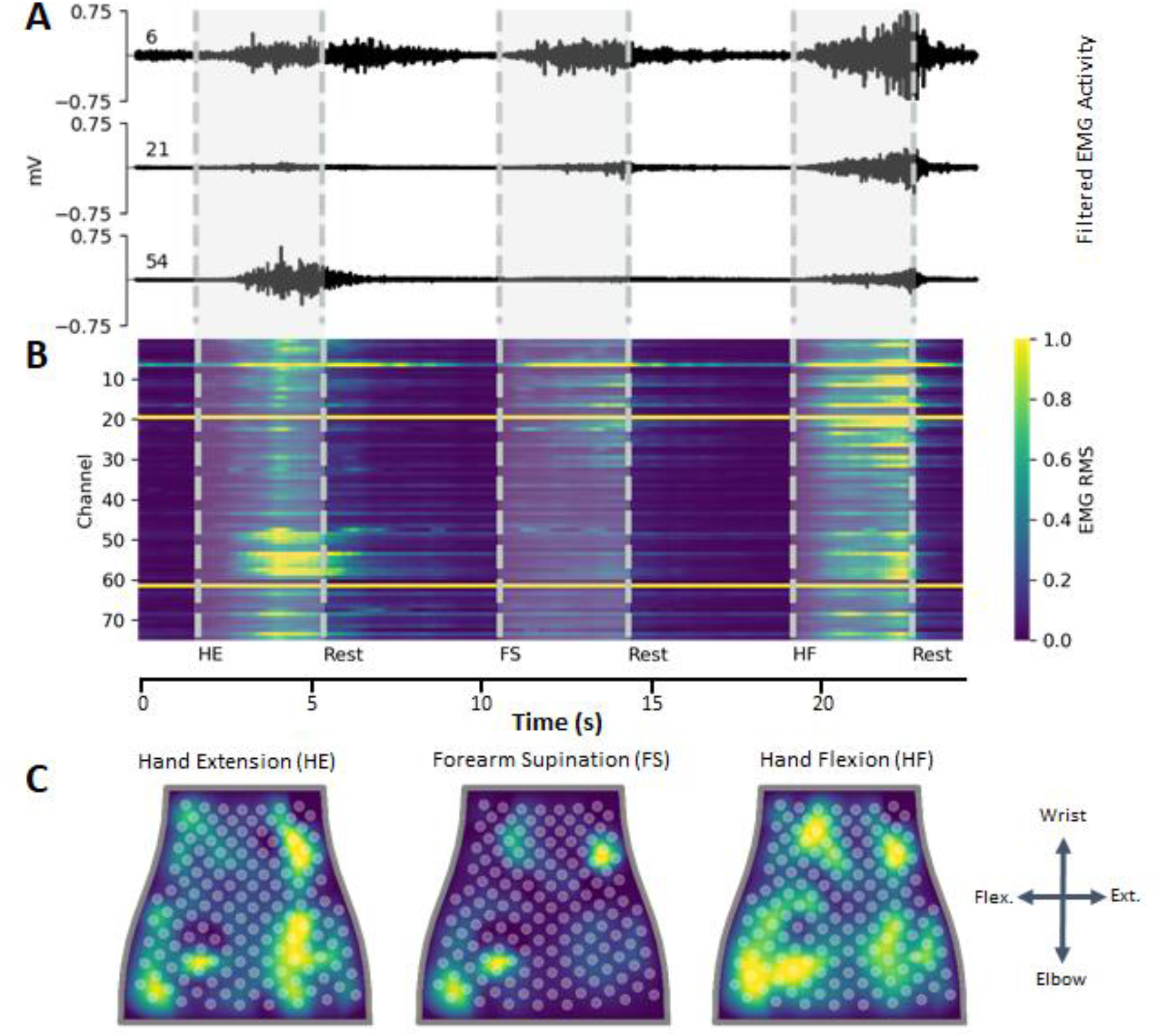
Representative EMG data recorded from participant with stroke. **(A)** Filtered EMG data recorded from 3 separate channels on the NeuroLife Sleeve during 3 movements: hand extension, forearm supination, and hand flexion. **(B)** Heatmap of normalized RMS activity, with the channel number on the y-axis and time on the x-axis. Note the activity across clusters of electrodes for each of the 3 separate movements. **(C)** Normalized RMS activity mapped to the sleeve orientation, with a legend showing the orientation of the sleeve mapping (flex. = flexors, ext. = extensors). Note the location of EMG activity is spatially located near the related musculature for each of the 3 movements.

Classification of movement intention in stroke participants was performed in two different ways: (1) using the 2.5s center during a cue or rest period, or (2) on the continuous timeseries data. For the 2.5s center window method, the middle 2.5s of each cue and rest period during a block was extracted (Figure 3A). This resulted in a total of 22 predictions of 100ms binned RMS data per cue (2.5s with the first three 100ms bins removed for containing out of window data at the beginning of the cue). This method was applied to both the training and testing datasets to reduce noise by removing the transition periods, similar to previous studies [21]. This dataset was used to evaluate different machine learning models for decoding the user’s movement intent. In able-bodied subjects, classification was performed as described above but using the 1.5s center during a cue or rest period, resulting in a total of 12 predictions. These data are presented in Figure 3 for participants with stroke, and Supplementary Figure 5 for able-bodied participants.

**Figure 3.**
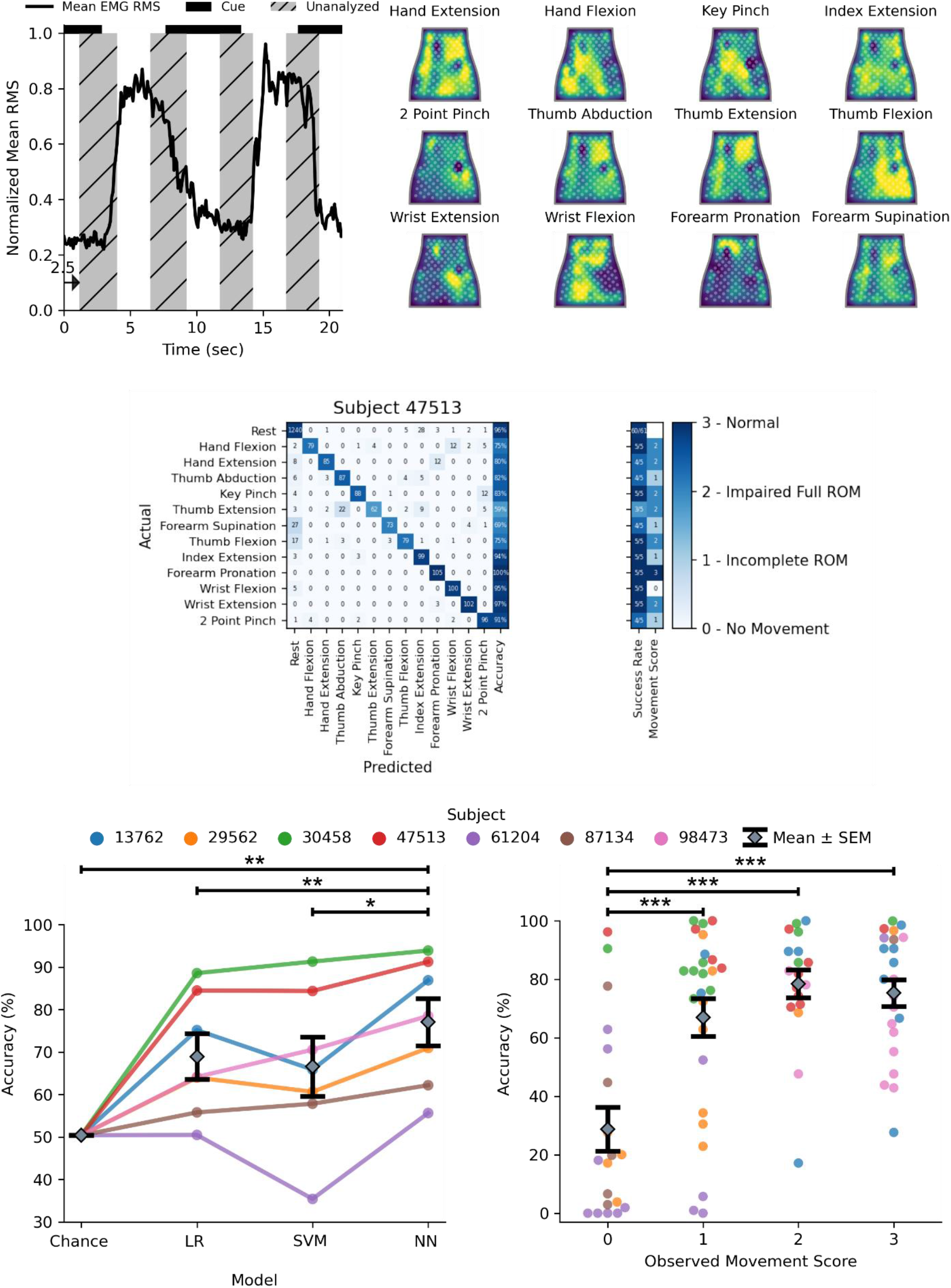
Decoding hand gestures using the NeuroLife EMG System. **(A)** Illustration depicting the data used for training and testing the decoder. The presentation of the cue is shown as a black bar on the top of the plot, and the middle 2.5 seconds of the cue presentation is used for analysis. **(B)** Heatmaps of various movements from a participant with stroke. **(C)** Decoding performance comparing 3 models: LR (Logistic Regression), SVM (Support Vector Machine), and NN (Neural Network). The NN outperforms both the LR and SVM models, as well as chance accuracy (Stroke Model: paired t-test NN vs. SVM, p=9.3 × 10^−3^; NN vs. LR, p=9.1 × 10^−4^; NN vs. Chance, p=4.4 × 10^−4^). **(D)** Association between the observed movement score and decoder performance of the neural network (Observed Movement Score, One-way ANOVA, Accuracy (%): F[3, 80] = 13.38, p= 3.7 × 10^−7^). The decoder struggles learning to predict movement attempts in which there was no observable movement (movement score = 0), and performs similarly when there is observable movement (movement score >= 1). **(E)** Confusion matrix for a participant with stroke detailing the decoding performance across all movements.

For decoding of continuous timeseries data, we performed a dynamic cue shifting technique to account for the variability in the participant’s ability to respond to the onset and offset of cues. Latency between cue onset and the onset of EMG activity is a persistent problem within decoding that can lead to significant deficits in algorithm performance and is exacerbated in data recorded from participants with neurological impairments such as stroke. Traditionally, these onset and offset variabilities are handled by shifting cues a predetermined amount of time based on reaction times [22], or can be assigned for each cue manually [18]. However, these methods still fail to capture the full distribution of onset and offset variability. Here we use an automated approach to dynamically shift cue labels to match the EMG activity. The average EMG signal was aligned with the intended cue times, and residuals were calculated between the EMG signal and the signal mean for each cue segment. The transition point between segments was then iteratively optimized to minimize the sum of squared residuals (Supplementary Figure 4). Cue timings were shifted up to a maximum time of 2s beyond the intended cue time. These data are presented in Figure 5.

### Classification

Classification was performed using all recording blocks (single and mixed). Importantly, the testing consisted of the final 4 recording blocks of data collected for that subject. In other words, none of the training set occurred later in time than the testing set to prevent data leakage of time dependent signal fluctuations that can significantly influence decoding performance.

Three classifiers were compared, including a logistic regression (LR) model, support vector machine (SVM), and a neural network (NN). For the LR and SVM models, data was additionally preprocessed using principal component analysis for dimensionality reduction, keeping components that accounted for >95% of the variance. LR and SVM models were trained using the scikit-learn toolbox [23] in Python 3.8. To optimize hyperparameters for both LR and SVM, a grid search on the training data with 5-fold cross validation was applied to tailor a specific model for each subject. Hyperparameter C was varied from 1e-4 to 1e4 for LR, and hyperparameters C and Gamma were varied from 1e-4 and 1e4 for SVM. The best performing model hyperparameter combinations for each were selected for evaluation.

The NN was developed in Python 3.8 using the FastAI package [24]. FastAI defaults were used for training except where noted. The model architecture takes an input of a flattened N channels x 4 array from the N channels of the sleeve and 4, 100ms windows of mean RMS signal. The input layer connects to two fully-connected dense layers, with size 1000 and 500 respectively, with batch normalization and the ReLU activation function between layers. The final layer had 13 classes corresponding to the 12 cued movements and rest. Finally, a Softmax activation function was applied to the model outputs to provide prediction probabilities for each of our movements. The predicted movement for a given time point was the movement with the greatest prediction probability. The training procedure used label smoothing cross entropy loss (p=0.9) and the Adam optimizer. During training, dropout was applied to each layer with 20% probability to prevent overfitting. The learning rate was optimized using the FastAI learning rate finder tool [24]. Each model was trained for 400 epochs with early stopping criterion, using the one cycle training policy from FastAI.

To measure performance, we use two complementary metrics. Accuracy is defined as the percentage of 100ms time bins predicted by the classifier to be the same as ground truth. Accuracy is a standard classification metric and provides a high temporal resolution metric of performance. We also present success rate as a decoding performance metric, similar to previous studies [22]. A movement is considered successful if there is at least 1s continuous period within a cue that is correctly decoded as the intended movement. The success rate is then calculated as the percentage of cues which are considered successful. This metric approximates an observer rating each cue as a binary success or failure and is more aligned with how a user would perceive performance.

All comparisons were planned in the experimental design *a priori*. Normality of distributions were tested using Lilliefors tests. Significant differences were determined using paired t-tests (Figure 3D, 4A) and unpaired t-tests where appropriate. Significant differences for multiple comparisons were determined using one-way ANOVAs followed by Tukey HSD tests (Figure 3D). Alpha of 0.05 were used for single comparisons. To correct for multiple comparisons, a Bonferroni-corrected alpha of 0.0167 was used for Figure 3C and an alpha of 0.025 was used for Figure 5A. Statistical tests for each comparison are noted in the text. Statistical analysis was performed in Python 3.8 using SciPy and Statsmodels. In all figures, * indicates p<0.05, ** indicates p<0.01, and *** indicates p<0.001. Error bars indicate mean ± SEM in all figures.

**Figure 4.**
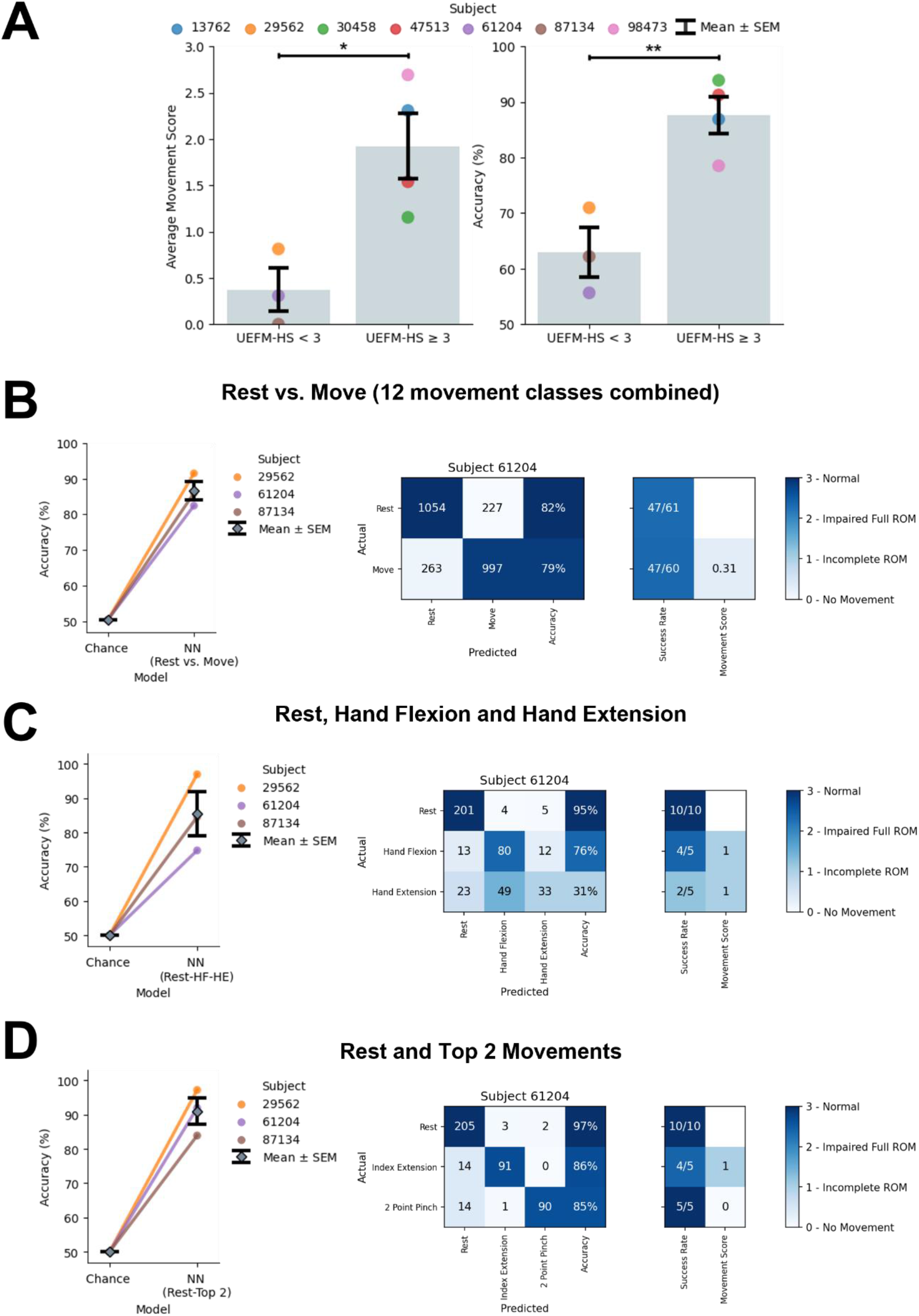
Decoding hand gestures in patients with severe hand impairment (UEFM-HS < 3). **(A)** *Left:* Comparison of severe (UEFM-HS < 3) and mild (UEFM-HS ≥ 3) subject impairment average movement scores (Average movement score: unpaired t-test UEFM-HS <3 vs. UEFM-HS ≥ 3, p=0.02). *Right:* Comparison of NN decoding performance for severe and mild subject impairments (Decoding accuracy: unpaired t-test UEFM-HS < 3 vs. UEFM-HS ≥ 3, p=0.006). **(B)** Decoding performance of NN binary classifier for UEFM-HS < 3 subjects comparing Rest and Move in which Move is made up of combining all 12 movements into a single class. Confusion matrix of subject 61204 for the two-class problem. The observed movement score is the average of all movement types’ observed movement scores. The two-class decoder can reliably distinguish the difference between a resting and moving state. **(C)**. Decoding performance of NN model when restricting classes to Rest, Hand Flexion, and Hand Extension. Confusion matrix of lowest performing subject (61204) for the three-class problem. The three-class decoder is not sufficient to distinguish the movements reliably. **(D)** Decoding performance of NN model when restricting classes to Rest and the top 2 movements for each subject for a total of three classes. Confusion matrix of subject 61204 for the three-class problem. Focusing on movements specific to subjects increases the robustness of decoder performance.

**Figure 5.**
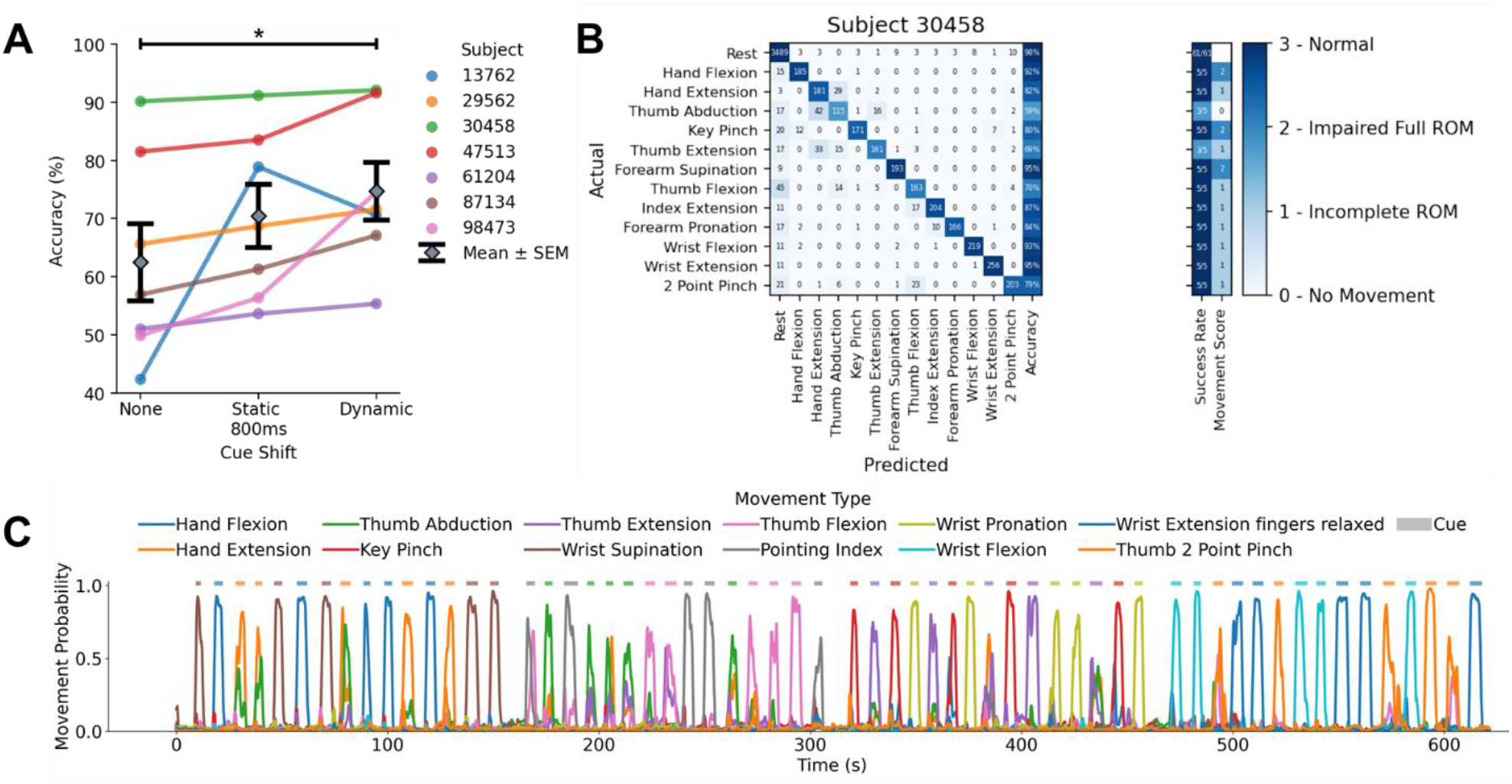
Decoding hand gestures in simulated real-time scenarios. **(A)** Dynamic cue shifting significantly improved accuracy compared to no cue shift (Cue shift: paired t-test Dynamic vs. None, p=0.020). There was no significant difference between a dynamic cue shift and static 800-millisecond cue shift (approximately the average cue shift across subjects) (Cue shift: paired t-test Dynamic vs. Static 800ms, p=0.22). **(B)** Confusion matrix detailing performance from one subject in the simulated real-time scenario. **(C)** Time series plot depicting decoder class probabilities across time. The presented cue is shown in above the time series plot as a rectangular colored bar with the color corresponding to the movement class.

## Results

### Movement intention can be inferred from forearm EMG activity of participants with stroke using the NeuroLife EMG System

To demonstrate the utility of the NeuroLife EMG System to sense and interpret muscle activity in the forearm, we first assessed the ability to decode hand, wrist, and forearm movement intention. Participants were guided through various blocks of movements and EMG data was recorded. To extract the most stable segment of the signal, the middle 2.5-seconds of each cue and rest period was isolated to remove transition periods (Figure 3A). By removing the transition periods and focusing on periods of consistent activity we are left with a standardized dataset to compare performance of various models.

Heatmaps of EMG activity across the sleeve are shown for one participant with stroke (Figure 3B). These heatmaps highlight the visual differences between forearm EMG activity across the various movements. EMG activity is less localized in the heatmaps of participants with stroke compared to the able-bodied participants (Supplementary Figure 2). This trend is consistent across stroke severity, with the more severely impaired participants having less localized forearm EMG activity (Supplementary Figure 9). These results are consistent with previous reports of lack of independent muscle control following stroke [25].

Decoding of EMG activity can be performed using a variety of different techniques, we therefore evaluated three commonly used machine learning decoding approaches including a logistic regression (LR) [26], support vector machine (SVM) [27], and neural network (NN) [28]. To validate our decoding pipeline, we tested decoding performance in able-bodied subjects across all 12 movements with the expectation of highly accurate decoding. Overall, the NN obtained 96.8±0.5% accuracy and outperformed LR and SVM models which had 91.5±0.8% and 90.8±1.2% accuracy respectively (Supplementary Figure 5; Able-Bodied Model: paired t-test NN vs. LR, p=5.8 × 10^−5^; NN vs. SVM, p= 1.6 × 10^−3^). These decoding results are consistent in the dataset comprised of participants with stroke attempting all 12 movements, where the NN obtained 77.1±5.6% accuracy, and outperforms the LR (69.0±5.4%) and SVM models (66.6±6.9%) (Figure 3C; Stroke Model: paired t-test NN vs. LR, p=9.1 × 10^−4^; NN vs. SVM, p=9.3 × 10^−3^). In summary, the NN outperforms the LR and SVM when decoding forearm EMG activity to infer movement intention. All subsequent analysis is performed in participants with stroke and using the NN for decoding.

Next, we investigated the relationship between the participant’s ability to perform a movement unassisted and our ability to accurately decode that movement. Generally, decoding performance improved as the observed movement score increased (Figure 3D; Observed Movement Score, One-way ANOVA, Accuracy (%): F[3, 80] = 13.38, p= 3.7 × 10^−7^). A comparison of decoding accuracy based on movement score was computed using a Tukey HSD test (Supplementary Table 2). For movements with visible motion (score >= 1), the overall decoding accuracy was 85.7±3.2% (Chance: 27.9%), whereas for movements where the participant had no visible motion (score=0) the accuracy dropped significantly to 27.3±3.2% (Chance: 4.0%) (Movement Ability: Movement score=0 vs. Movement score=1-3: unpaired t-test, p=3.9 × 10^−9^).

Next, we investigated decoding performance of individual movements in participants with stroke. The confusion matrix with individual movements for one subject is shown in Figure 3E. The best performing movements across subjects were Rest, Wrist Flexion, and Index Extension with an average accuracy of 93.3±6.5% (Figure 3E, right column). On average across subjects, the worst performing movements were Forearm Supination and Thumb Abduction, with an average accuracy of 39.4±9.9% (Supplementary Figure 6). The success rate per movement type for one subject is presented in the right column of the confusion matrix (Figure 3E). Overall grand average success rate across all movements achieved 75.9±4.2%. The top three movements had an average success rate of 93.2±7.6% (Successes/Attempts; Rest: 409/425, Index Extension: 23/30, Wrist Flexion: 20/30), with the bottom two movements obtaining a much lower success rate of 36.0±11.5 (Successes/Attempts; Forearm Supination: 14/43, Thumb Abduction: 17/43).

### Decoding movement subsets to achieve high performance in participants with severe stroke impairments

As the decoding performance of our algorithms was dependent on the presence of visible movement in our participants, we next investigated the association of hand impairment severity based on the Upper Extremity Fugl-Meyer Hand Subscore (UEFM-HS) with observed movement scores and decoding performance (Figure 4A). Both the observed movement score and decoding performance in participants with severe hand impairment (UEFM-HS < 3) is significantly different than in individuals with moderate or mild hand impairment (UEFM-HS ≥ 3) (Average movement score: unpaired t-test UEFM-HS < 3 vs. UEFM-HS ≥ 3, p=0.02; Decoding accuracy: unpaired t-test UEFM-HS < 3 vs. UEFM-HS ≥ 3, p=0.006). Our complete 12-movement survey is helpful for understanding what movements may be decodable for each participant and may be appropriate for facilitating ATs in individuals with moderate or mild hand impairments. However, those with severe hand impairments are unlikely to be able to accurately control that many movements. Instead, it may be desirable to subset down to a smaller number of movements, customized to the individual, that they can accurately control.

First, we assessed if sufficient signal was present to decode general muscular activity during cued movement periods compared to rest. Practically, this decoding scheme would enable an individual with severe hand impairment to control an AT with a single movement. We separated the problem into two classes (Rest vs. Move) where the “Move” class consists of combining the 12 different movements into one class (Figure 4B). The NN decoder was able to achieve high performance in individuals with severe hand impairment with 86.7±2.6% accuracy and 85.2±3.6% success rate (Successes/Attempts; Rest: 164/185, Move: 151/185). These results indicate that the surface EMG collected from individuals with severe hand impairment is sufficient for binary scenarios.

Encouraged by the binary decoder performance, we extended our analysis to include key functional movements for restoring grasp function, namely Rest, Hand Flexion, and Hand Extension (Figure 4C). With these key movements in individuals with severe hand impairment, the decoding performance achieved 85.4±6.4% accuracy and 88.0±7.7% success rate (Successes/Attempts; Rest: 45/46, Hand Flexion: 22/23, Hand Extension: 14/23). While, decoding the movements to enable hand flexion and hand extension is ideal for intuitive control of an AT, alternatively decoded movements with the greatest performance can be mapped to the most impactful functional movements. Thus, we tested decoding only the top performing movements for each subject (Figure 4D). When comparing Rest and the top two movements for each individual, decoding performance achieved 91.0±3.9% with grand average success rate of 90.6±4.2% (see Supplementary Table 4 for full details). This performance is comparable to the decoding performance of individuals with UEFM-HS ≥ 3 on 12 movements (87.6±3.4%) and provides a reasonable alternative for participants with more severe impairments.

### Decoding continuous forearm EMG data in simulated real-time scenarios in chronic stroke survivors

To demonstrate the utility of the NeuroLife EMG System to interpret muscle activity from the forearm to act as a control signal for assistive devices, we next tested our decoding algorithms in simulated real-time scenarios. Following a stroke, the ability to contract and relax muscle groups is slowed and highly variable [29], which consequently makes automated labeling of cues using a static time shift (e.g., 800ms) for training machine learning models imprecise. To account for this cue onset and offset variability, we first performed a dynamic cue shifting technique to automatically shift cue labels to match EMG activity (Supplementary Figure 4A). An average of 843±95ms of cue data per cue change or a grand average of 16.1±1.0% of the full cue data stream across all subjects was shifted using this technique (Supplementary Figure 4B). To verify this method, we compared decoding performance with and without cue shifting. Dynamic cue shifting significantly improved decoding performance achieving 74.7±5.0% overall with no cue shift achieving 62.5±6.7% (Figure 5A; Cue Shift: paired t-test Dynamic vs. None, p=0.020). Comparing dynamic cue shifting with a static 800ms shift (70.5±5.4% decoding accuracy) representing an estimate of the average dynamic shift, we determined no significant difference (Figure 5A; Cue Shift: paired t-test Dynamic vs. Static 800ms, p=0.22). One subject (13762) had an increase in decoding performance from a static shift with the rest of the subjects experiencing a decrease or no change in performance. Therefore, a dynamic cue shifting technique may present a more robust and automated solution to account for differences in subjects. Improvement in decoding performance from using dynamic cue shifting is likely due to 1) improved accuracy of the timing of cue onset and offsets in the training data which gives a better representation of each movement and thus better decoding performance, and 2) more accurate testing alignment and better testing parameters. These results suggest that cue labeling can substantially affect overall decoding performance in real-time decoders, and intelligent cue labeling can improve overall performance.

Using the dynamic cue shifting technique, we investigated decoding performance of individual movements in the simulated real-time dataset. The confusion matrix with individual movements for a single subject is shown in Figure 5B. The best performing movements across subjects were Rest, Wrist Flexion, and Wrist Extension, with an average accuracy of 89.7±7.3% for these movements. The worst performing movements across subjects were Forearm Supination and Thumb Abduction, with an average accuracy of 29.5±9.0%. A continuous time series plot of movement probabilities is shown in Figure 5D. Shaded regions indicate the cued movement with the probability of the movement type decoded based on motor intention.

To assess whether the NN decoder could be used in real-time situations, inference testing was conducted using a Surface Book 2 with NVIDIA GeForce GTX 1060 GPU. The trained NN decoder was exported and loaded in using the Open Neural Network Exchange (ONNX) Runtime [30] for inference testing. NN forward model prediction times on average took less than 1ms (307±49µs). Taking the entire preprocessing pipeline into consideration in addition to the NN forward prediction, the total inference time was 23.1±4.4ms. Since the resulting inference time is under 100ms (time bin for RMS feature calculation), the NN model is suitable for real-time inference.

### The NeuroLife Sleeve meets usability needs of chronic stroke survivors

Usability is a critical factor in the long-term adoption of an AT. Inconveniences of setup and comfort, as well as frustrations with reliability can often lead to eventual device abandonment. Therefore, in our final EMG recording session with each participant, we collected initial usability data of the NeuroLife Sleeve for use in chronic stroke survivors to help guide future development efforts. The questions posed to participants here were adapted to investigate overarching themes mentioned by stroke survivors, caregivers, and HCPs for the use of an assistive technology [4]. Participants answered each question on a 1 to 5 scale, and questions were targeted at the following categories: simple to apply, comfort for long-term use, freedom of movement during use, functionality / lightweightness and portability, potential for clinical and home use, and overall aesthetic design of the device (Supplementary Table 5). Summary data from the usability questionnaire is presented in Figure 6. For usability metrics with more than one question (e.g. simple to apply), the mean value was scored for that assessment.

**Figure 6.**
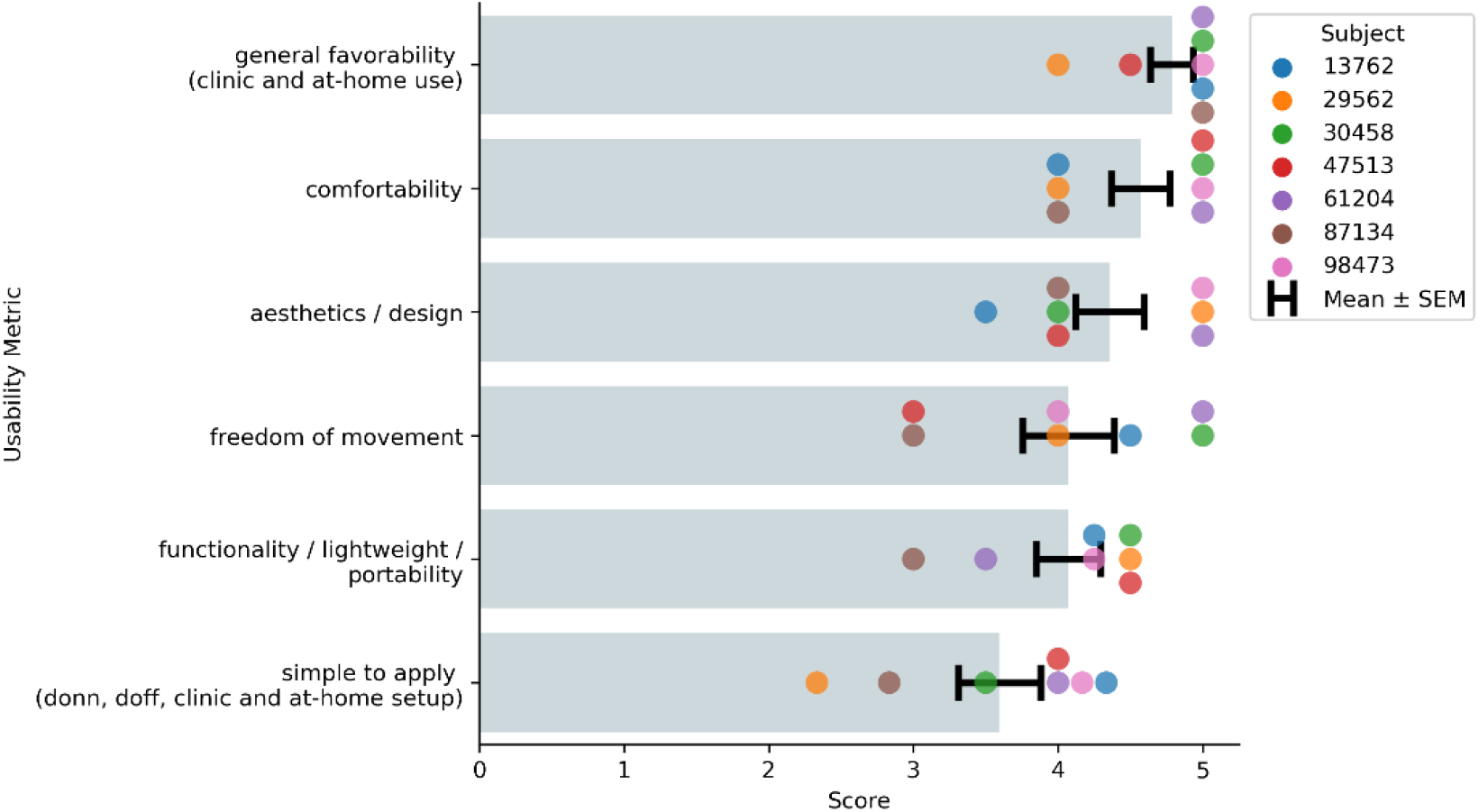
Summary of the NeuroLife Sleeve usability data from participants with stroke. Each participant with stroke ranked the NeuroLife Sleeve based on 6 usability domains. Group data is presented for each of the 6 domains.

In general, participants were optimistic that they could don and doff the NeuroLife Sleeve with the help of a caretaker in their home (3.60±0.28). Concerns were generally centered around the pre-application of the conductive spray and relative positioning of the system, which we are actively addressing in our next design iteration. During sessions, participants had the sleeve donned for >1.5 hours, and all participants reported general satisfaction with the overall comfort of the device (4.57±0.20). ATs should not hinder movement so that the user can successfully perform rehabilitation exercises or complete normal daily activities while using the device. The sleeve was designed with a lightweight stretchable fabric, and participants were generally satisfied with the ability to move their arm while the sleeve was donned (4.07±0.32). Participants were highly confident (4.07±0.22) that they could wear the sleeve doing functional light activities around their home, suggesting that the sleeve is non-restrictive, lightweight, portable, and promising for home use. A commonly overlooked barrier to widespread adoption of assistive technologies is user acceptance of the overall look and feel of the device [4]. Overall, all participants were extremely satisfied with the overall design of the sleeve (4.36±0.24). In general, they were all very excited for the opportunity to use the sleeve with the “general favorability” metric receiving the highest score of 4.79±0.15. In summary, the usability results from the current study provide promising early data that the NeuroLife Sleeve can meet end user needs with directions on where to improve for future iterations.

## Discussion

In this study, we demonstrate decoding of motor intention using the NeuroLife EMG System in people with upper-limb hemiparesis due to chronic stroke. Based on high-density surface EMG data collected from the forearm,12 functional hand, wrist and forearm movements were classified with high accuracy. Overall decoding accuracy was associated with the participant’s ability to perform the movement (quantified here as observed movement score), with greater functional movement corresponding with higher decoding accuracy. Even in movements with little to no movement capacity (movement score <= 1), the system was able to accurately differentiate movement intent, albeit with some decrease in performance. This demonstrates the NeuroLife EMG System’s ability to infer movement intention in stroke survivors with severe motor impairments.

Previous studies have demonstrated decoding of motor intention using surface EMG in the upper extremity in chronic stroke survivors [18,31–33]. In these studies, a range of machine learning techniques, impairment levels of the participants with stroke, and types of movements have been investigated. Classification accuracy was comparable to previous work with similar movement sets, although differences in study methodology restrict direct comparison. Of note, recent studies have shown encouraging results with a limited set of manually placed electrodes, which may account for some performance differences [18,32]. Moreover, localizing electrodes to muscular activity critical to grasp production can be an effective strategy to minimize system complexity. The optimization of electrode placement and reduction of hardware complexity is a planned future direction for the NeuroLife Sleeve. Additional studies have used similar numbers of channels as we have presented [18,34,35]. However, the systems used in the previous studies were laboratory grade EMG systems that require lengthy and difficult setup processes by trained technicians. Usability around donning/doffing is a key concern for adoption of AT, and systems with extensive setup procedures risk poor acceptance in clinics, rehabilitation settings, and the home. Prior studies have also shown that time domain features, such as RMS, combined with NN approaches can outperform more classical statistical or machine learning approaches. Our results agree with these findings, further supporting that high density EMG recordings have sufficient complexity to leverage the recent developments in deep learning. We extend the findings of previous studies by presenting an easy to don and doff wearable device that removes the need for manual placement of electrodes. This reduces the necessary setup time and ensures consistent placement of recording electrodes across sessions. Additionally, we present data to support the real time performance of the decoding paradigm. Our device can decode motor intention with high performance across a variety of subjects, with fast enough speed to reliably perform inference alongside data collection. Finally, we present a viable, automated cue shifting method that removes the necessity for manual relabeling and improves system performance.

Usability is an important factor for clinical technologies to assist with stroke rehabilitation by supporting motivation for consistent and active training. While existing AT solutions show promising results, these systems tend to focus on the technology and often fall short in the user-centric designs. Most clinical ATs involve manual placement of patch electrodes and long calibration procedures which limits the amount of practice that can be achieved within a given rehabilitation session. Furthermore, many systems are bulky and lack portability, which can limit patient adoption for use outside of rehabilitation training and into the home [36]. Here, we demonstrate that the NeuroLife EMG System can address many usability concerns of current technologies while providing robust decoding of motor intention. In combination with soft exoskeletons or FES, the sleeve can drive intention-based training coupled with functional movements in a user-centric form factor.

We collected and report user feedback to quantify features of the system that end-users were satisfied with, and identify areas for further development efforts. Based on user feedback from the current study, the sleeve design meets various end user needs. The design allows for use on either arm, and the stretchable, lightweight fabric design was reported by participants to be comfortable and does not limit natural arm movements. Aesthetically, subjects were pleased with the sleeve design and advocated that they would use the system at home for rehabilitation and activities of daily living given the opportunity. Participants mostly agreed that the sleeve was straightforward to don and doff during the study with the help of the researchers, and believed that they could apply the sleeve with the help of a caretaker. However, participants identified the simplicity to apply the sleeve as an area that is currently lacking, and participants were not confident in being able to apply the sleeve independently without assistance. This is an identified area for future development and will be the focus of next design iterations to enable at-home use. Despite this current usability limitation, participants indicated that not only would they feel comfortable performing rehabilitation therapy at home, but are excited for the possibility of using the sleeve as a therapy tool indicated by the highest score for general favorability.

The present study provides an initial demonstration of the NeuroLife EMG System to decode motor intention in chronic stroke survivors while simultaneously meeting needs, but some limitations merit consideration. While the reported results indicate that the Neurolife EMG System can be used to decode motor intention in a package that meets end user needs, there is still room for improvement in various areas including refinement of decoding algorithms, the sleeve design and related hardware, and eventual applications. Future work refining decoding algorithms will focus on overall improvements to decoding performance by leveraging many of the advancements made in recent years in the field of deep learning. We will investigate the use of more complex neural network models, including recurrent neural networks (RNNs) and transformers optimized for time series modeling and which could improve overall decoding accuracy, specifically for participants with limited movement capability. Additionally, we will apply various machine learning techniques including unsupervised learning to expedite setup and calibration times for new users to address this important aspect of usability. Improvements to data quality itself can be accomplished with visual reinforcement to subjects. An online decoding system that displays the decoded intention may be more beneficial to participant engagement over the image cues used in the current study. While we provide the initial proof-of-concept demonstration of the NeuroLife EMG System here, the data collected during the study was not representative of how the system will be ultimately deployed as an assistive device. For example, in the current study participants kept their elbow stationary on the table during movements and did not interact with objects, both of which can significantly influence forearm EMG activity and thus decoding performance. Future studies will focus on capturing training data in more complex situations, such as during reach and grasp tasks and object manipulations, to develop decoders robust to movement. Similarly, the decoding performance presented here was in the absence of assistive device control. Commonly used assistive devices including FES and exoskeletons may interfere with EMG activity when active and thus can significantly affect decoding performance [31,37]. Our group is working to integrate FES functionality within the same EMG recording electrodes to eliminate the need for additional hardware such as an exoskeleton or additional patch electrodes. Future work from our group will focus on developing algorithms that can decode EMG during FES activity. With a technology that incorporates EMG and FES into a single consolidated sleeve, the system has the potential to help support motor recovery and assist in ADLs [14,22].

## Conclusion

The focus of this study was to validate the NeuroLife EMG System by decoding hand, wrist, and forearm movements and collect usability data from participants with stroke. We demonstrate accurate EMG decoding of 12 different movement classes with a neural network in both able-bodied and stroke participants. Decoding accuracy was associated with the movement ability of the participant. The decoding results are consistent with similar myoelectric intention-based studies. Finally, we present data on the common usability factors of assistive devices including the simplicity, comfortability, portability, and weight of the sleeve. Overall, all participants reported good to outstanding ratings for each of the usability categories, indicating that the NeuroLife EMG System can provide accurate decoding of upper extremity motor intention while meeting the usability needs of end users.

## Data Availability

Data will be made available upon request.

## Acknowledgements

The authors would like to thank our development and management teams at Battelle Memorial Institute including Jesse Keckler, Nick Annetta, Sam Colachis, and Josh Branch for their engineering contributions, Charli Hooper for her assistance with data collection, and Andrew Sweeney for his contribution to the manuscript graphics. Financial support for this study came from Battelle Memorial Institute.

## Data Availability

The data that support the findings of this study are available upon reasonable request from the authors.

## Supplementary Material

### Supplementary Methods

#### Inclusion and Exclusion Criteria

For physically impaired individuals, inclusion criteria address the minimum length of time since the stroke that led to the impairment. Inclusion criteria may also pertain to meeting dimensional requirements related to interacting with the system hardware (e.g., subject’s arm dimensions must be such that they can appropriately don an existing electrode sleeve design).

For populations with potential for cognitive impairment (e.g., stroke survivors), inclusion criteria indicating ability to follow 3-step commands and communicate verbally (e.g., at least able to provide yes/no responses with accuracy) apply.

Specific Inclusion Criteria include:

1. Males and females ≥ 18 years old
2. Chronic stroke survivors who are at least 180 days post-stroke
3. Ability to provide appropriate consent to partake in the study
4. Ability to follow 3-step commands and deemed by an occupational therapist to have the capacity to complete required upper extremity movements
5. Ability to secure transportation to attend scheduled study sessions
6. Stroke-related hand impairment that interferes with ability to complete activities of daily living and is classified as Stage 1-6 on the hand subscale of the Chedoke McMaster Stroke Assessment

Persons with life-supporting or sustaining equipment or critical non-removeable implanted electronic devices are excluded for safety reasons since it is not known if the experimental systems would interfere with this equipment.

Specific exclusion criteria include:

1. Presence of any other clinically significant medical comorbidity for which, in the judgment of the Investigator, participation in the study would pose a safety risk to the subject
2. Currently participating in physical rehabilitation (e.g., occupational or physical therapy) for stroke-related upper limb impairment
3. Co-occurring neurological condition (e.g., Parkinson’s disease, Multiple Sclerosis) or other neuromuscular disorder (e.g., Carpal Tunnel Syndrome, neuropathy) that, in the judgment of the Investigator, may influence study results
4. Individuals who are immunosuppressed, have conditions that typically result in becoming immunocompromised, taking chronic steroids, or currently receiving immunosuppressive therapy
5. Individuals having or requiring any of the following: implanted pacemaker, life supporting/sustaining equipment, or critical non-removable implantable electronic devices such as an insulin pump or neurostimulator. An implanted Medtronic LINQ monitor does not meet this criterion (i.e., patients with a LINQ monitor may participate in this study).
6. Persistent pain ≥ 7/10 in impaired upper extremity, as measured by Numeric Pain Rating Scale (0-10)
7. Individuals whose forearm is determined to be too small or too large to fit the electrode sleeve being investigated.
8. Individuals who are pregnant or plan to get pregnant during the course of the study (self-report).

## Supplementary Figures

**Supplementary Figure 1.**
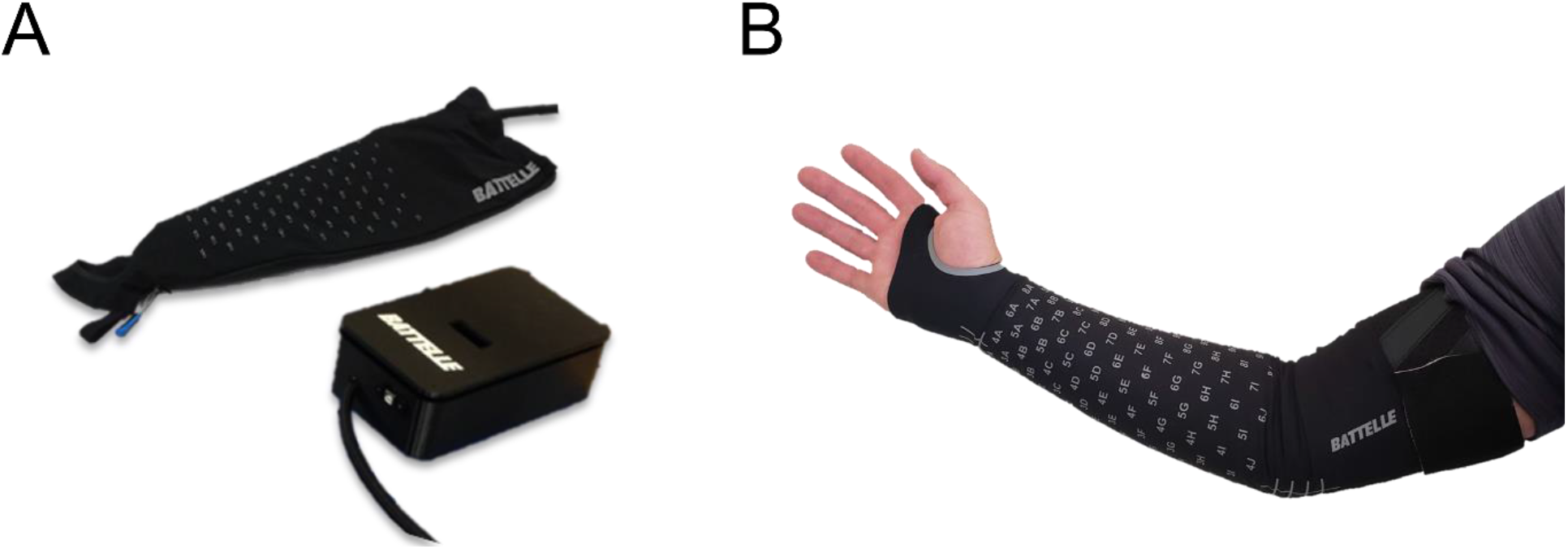
**(A)** Configuration of hardware used for EMG data collection showing the sleeve and EMG signal acquisition module (ESAM). **(B)** View of the sleeve donned on a participant.

**Supplementary Figure 2.**
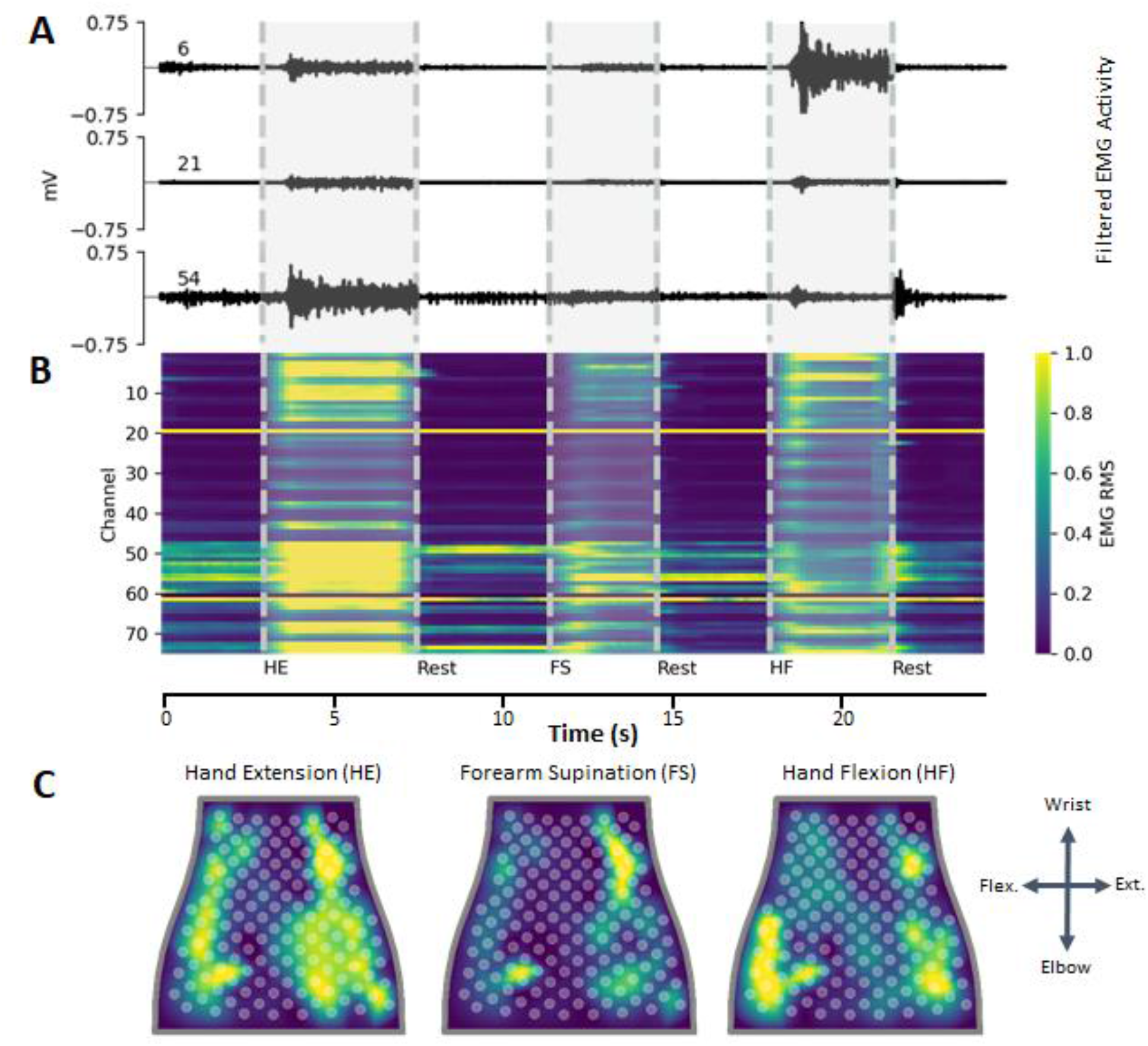
Representative EMG data recorded from able-bodied participant. **(A)** Filtered EMG data recorded from 3 separate channels on the NeuroLife Sleeve during 3 movements: hand extension, forearm supination, and hand flexion. **(B)** Heatmap of normalized RMS activity, with the channel number on the y-axis and time on the x-axis. Note the activity across clusters of electrodes for each of the 3 separate movements. **(C)** Normalized RMS activity mapped to the sleeve orientation, with a legend showing the orientation of the sleeve mapping (flex. = flexors, ext. = extensors). Note the location of EMG activity is spatially located near the related musculature for each of the 3 movements.

**Supplementary Figure 3.**
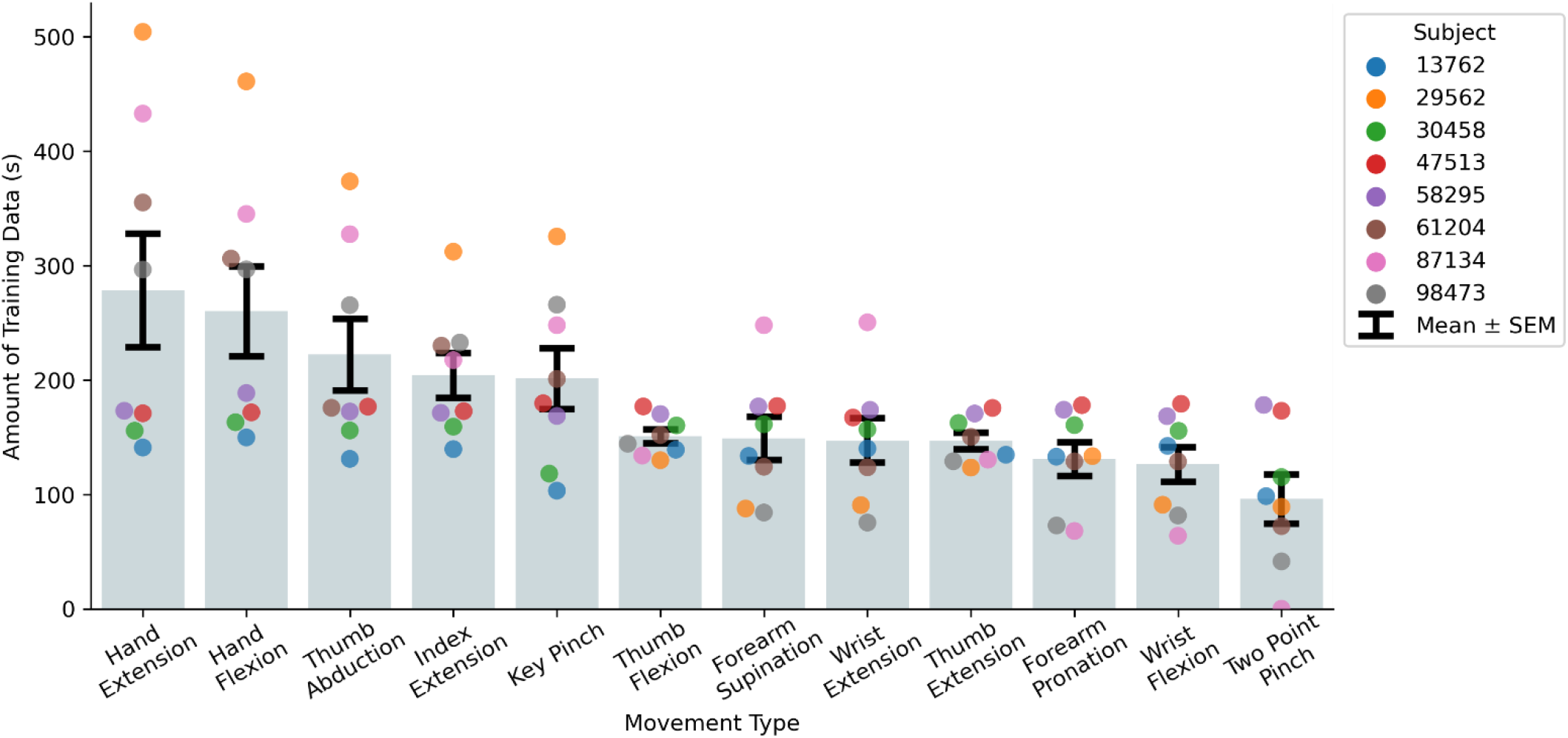
Amount of EMG data collected in time for each movement type per subject to train the decoders.

**Supplementary Figure 4.**
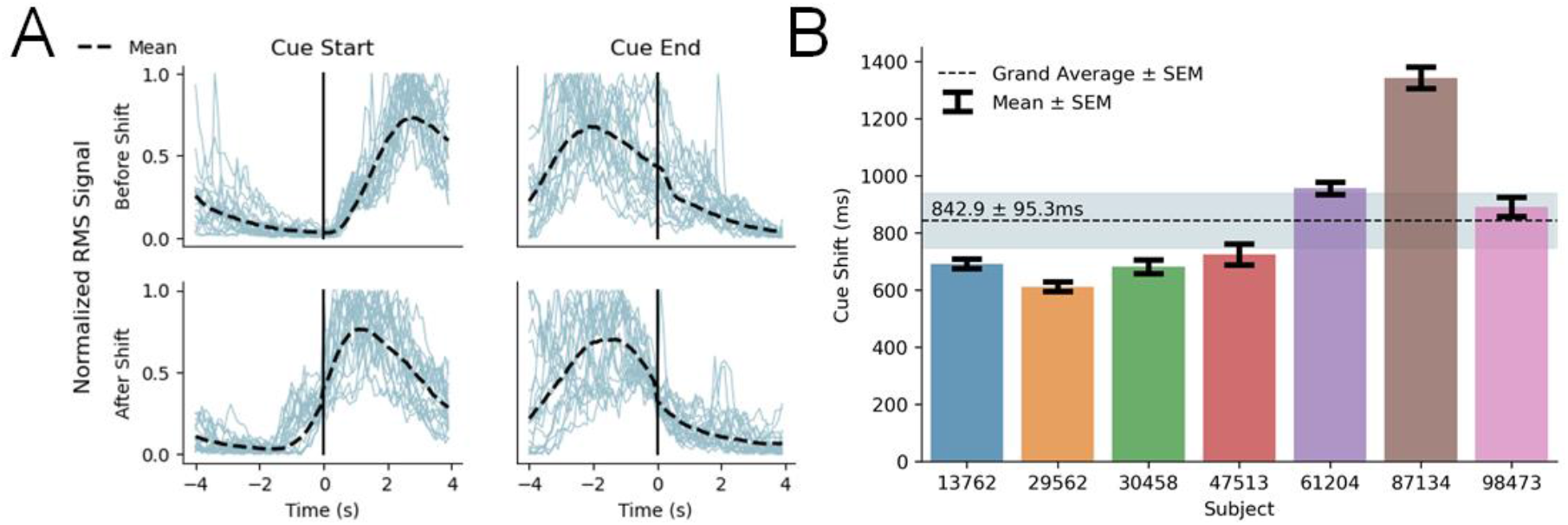
Dynamic cue shifting. **(A)** Cue shifting to improve the alignment of RMS of EMG signals during cue start and end times. Without dynamic cue shifting, the detected EMG signal is delayed during cue onset due to reaction time deficits and is sustained through the end of the cue due to residual muscular activity. By shifting dynamically based on underlying EMG activity, we properly align the intended cue with motor intent during training for better decoding performance. **(B)** Average cue shift per subject determined from the dynamic cue shifting technique.

**Supplementary Figure 5.**
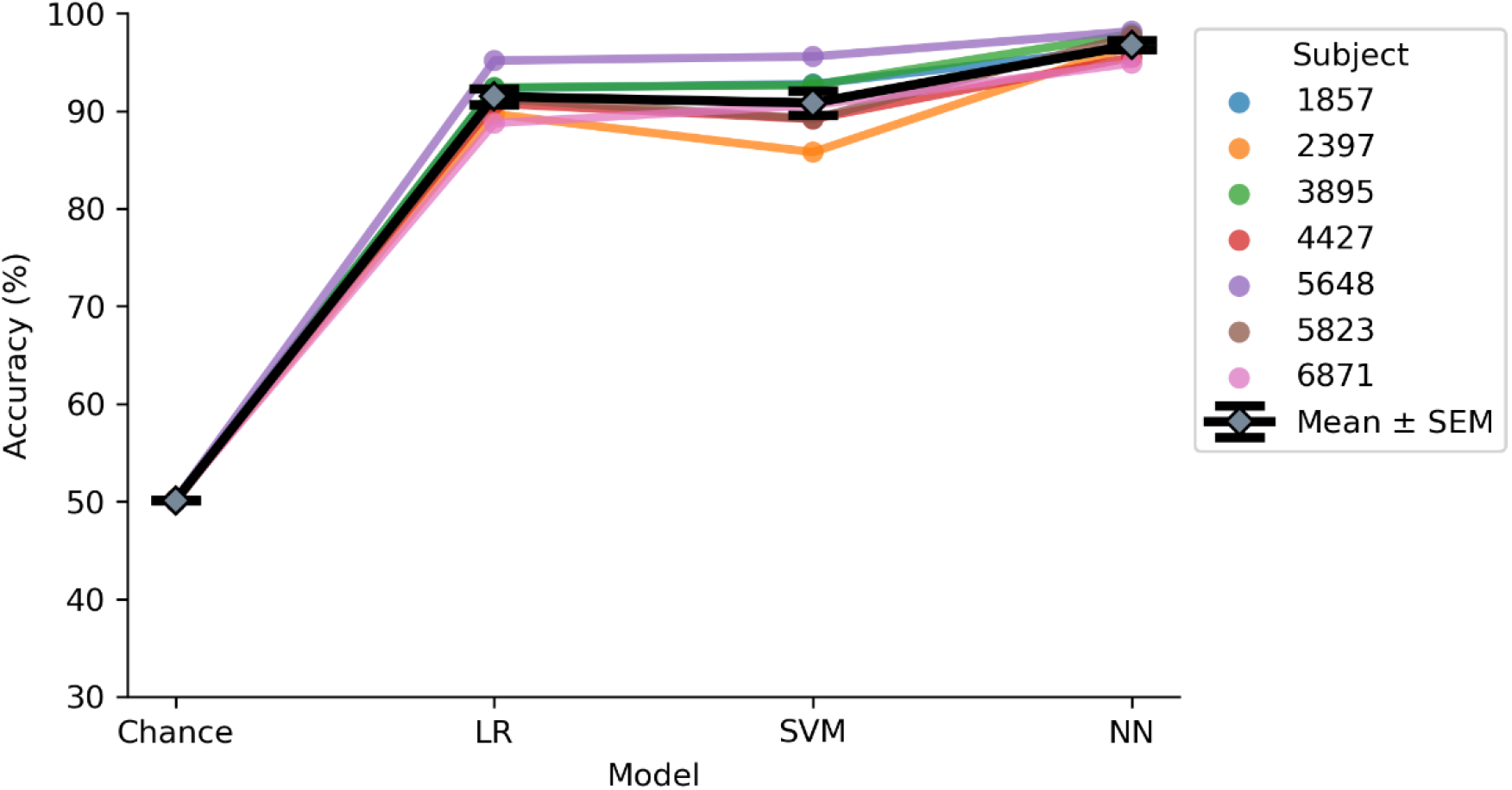
Model performance for able-bodied participants comparing 3 models: LR (Logistic Regression), SVM (Support Vector Machine), and NN (Neural Network). The NN outperforms both the LR and SVM models (Model: paired t-test NN vs. LR, p=5.8 × 10^−5^; NN vs. SVM, p= 1.6 × 10^−3^).

**Supplementary Figure 6.**
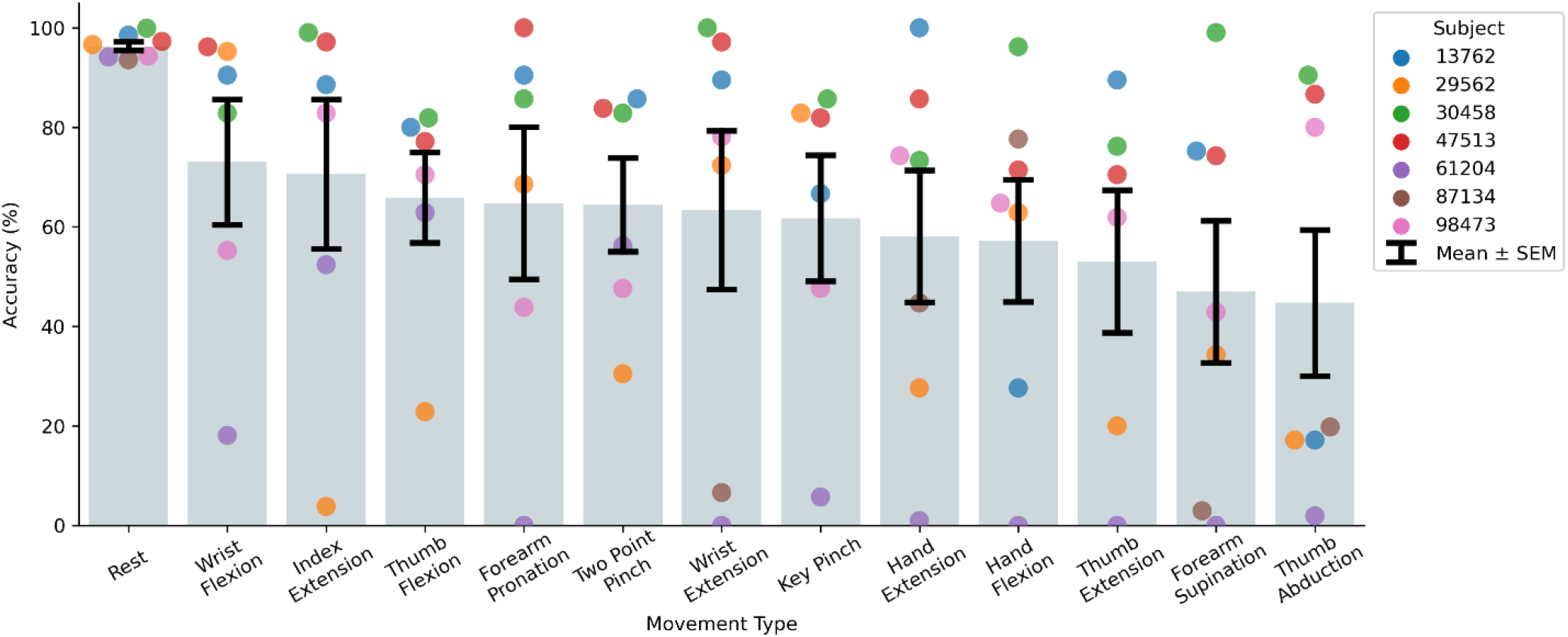
Decoding performance of the NN model based on movement type per subject.

**Supplementary Figure 7.**
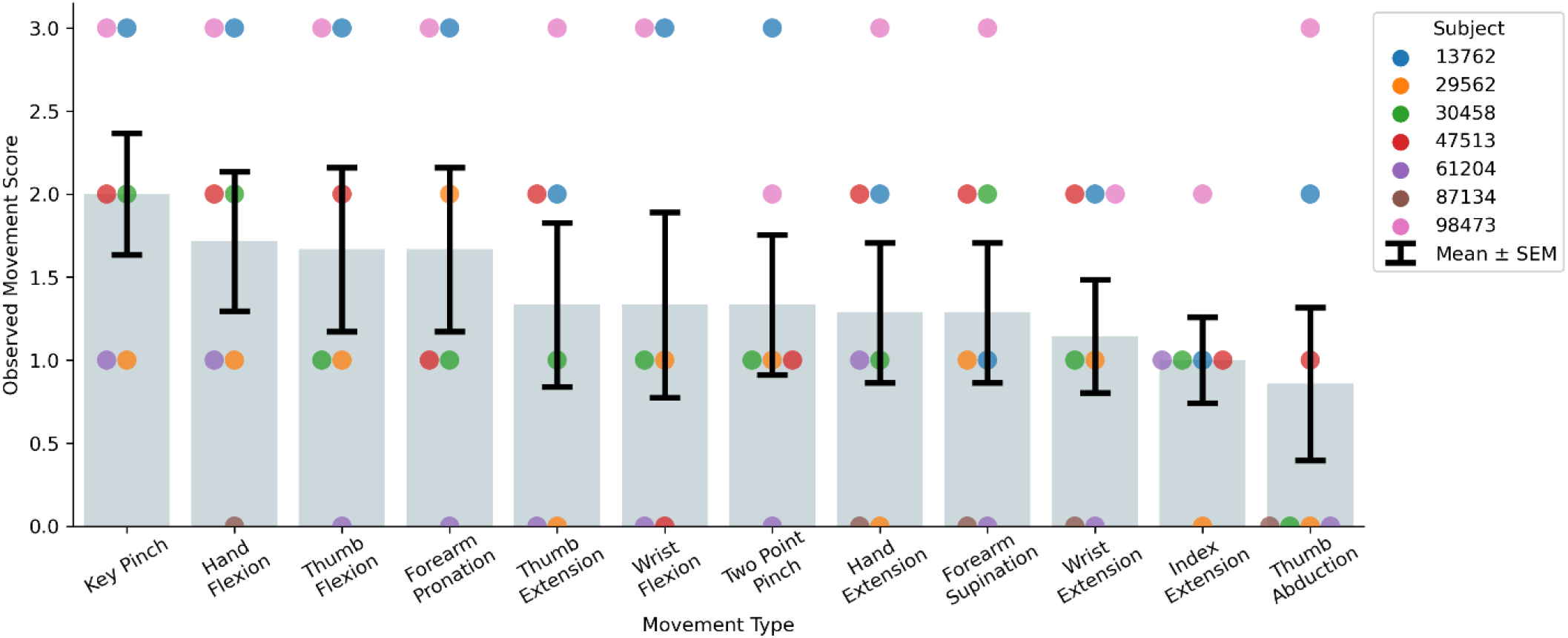
Observed movement score for each of the functional movements per subject ranked in order of ability from left to right. The Key Pinch and Hand Flexion were the simplest for subjects to perform whereas more complex movements such as Index Extension and Thumb Abduction were more challenging for stroke participants.

**Supplementary Figure 8.**
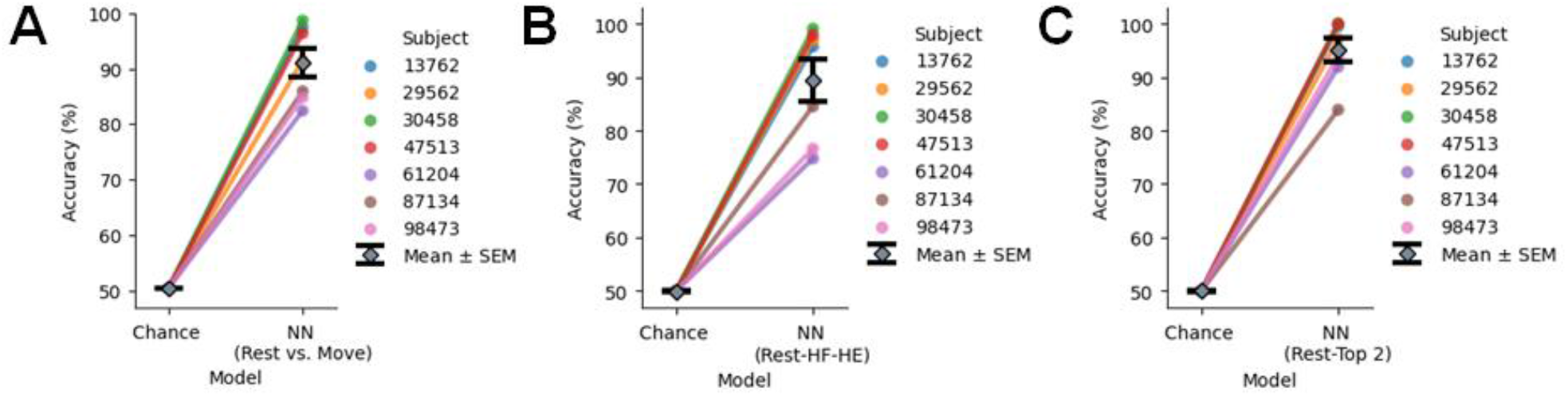
Decoding performance of NN model when down-selecting classes across all subjects. **A)** Decoding performance of NN binary classifier comparing Rest and Move in which Move is made up of combining all 12 movements into a single class. **(B)** Decoding performance of NN model when restricting classes to Rest, Hand Flexion, and Hand Extension. **(C)** Decoding performance of NN model when restricting classes to Rest and the top 2 movements for each subject for a total of three classes.

**Supplementary Figure 9.**
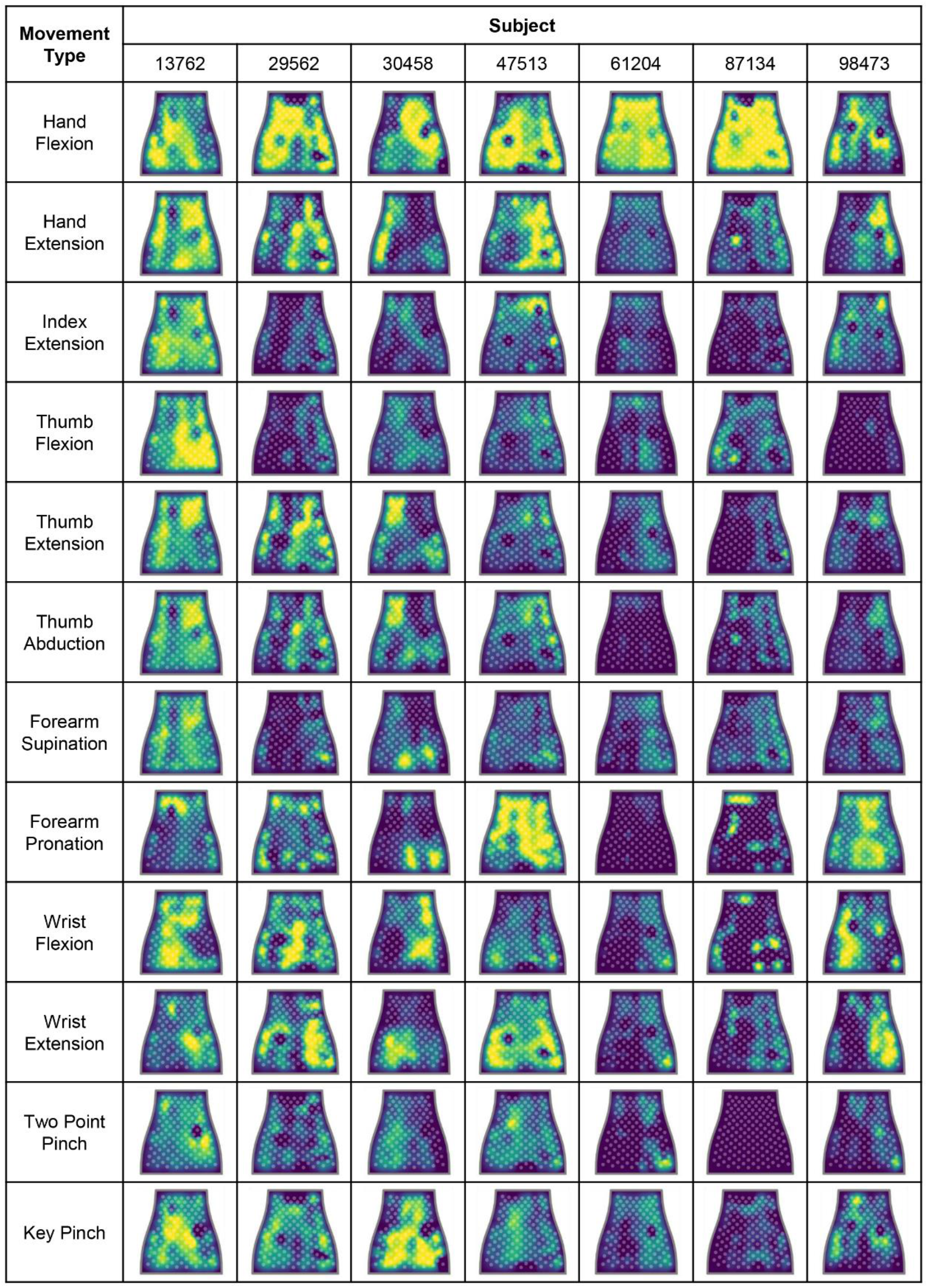
Normalized RMS activity mapped to the sleeve across all subjects and movements. Refer to Figure 2 for sleeve heatmap orientation.

**Supplementary Table 1.** Characteristics of the 3 different sized NeuroLife Sleeve.

|  | Small | Medium | Large |
| --- | --- | --- | --- |
| Number of Electrodes | 128 | 142 | 150 |
| Number of Electrode Pairs | 64 | 71 | 75 |
| Weight (grams) | 180 | 195 | 220 |

**Supplementary Table 2.** Demographics of able-bodied participants.

| Participant | Age Range, years | Sex |
| --- | --- | --- |
| 1857 | 26-30 | Male |
| 2397 | 31-35 | Female |
| 3895 | 26-30 | Female |
| 4427 | 26-30 | Male |
| 5648 | 21-25 | Female |
| 5823 | 21-25 | Female |
| 6871 | 26-30 | Male |

**Supplementary Table 3.** Tukey HSD multiple comparison results of decoded accuracy vs. movement ability. Subjects not able to perform the movement at all (Movement score=0) had a statistically significant difference in decoding performance compared to movements with scores ≥ 1.

| Group 1 Movement Score | Group 2 Movement Score | Mean Accuracy Difference (%) | P-value | Reject |
| --- | --- | --- | --- | --- |
| 0 | 1 | 38.2 | 0.001 | True |
| 0 | 2 | 49.7322 | 0.001 | True |
| 0 | 3 | 46.5521 | 0.001 | True |
| 1 | 2 | 11.5323 | 0.5262 | False |
| 1 | 3 | 8.3521 | 0.703 | False |
| 2 | 3 | -3.1802 | 0.9 | False |

**Supplementary Table 4.** Success rate of top performing movements in participants with severe hand impairment (UEFM-HS < 3).

| Subject | Top Movements: Successes/Attempts |  |  |
| --- | --- | --- | --- |
| 29562 | Rest: 10/10 | Key Pinch: 5/5 | Wrist Flexion: 5/5 |
| 61204 | Rest: 10/10 | Two Point Pinch: 5/5 | Index Extension: 4/5 |
| 87134 | Rest: 24/26 | Hand Flexion: 11/13 | Hand Extension: 7/13 |

**Supplementary Table 5.**
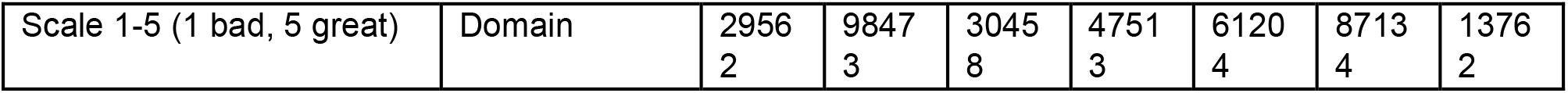
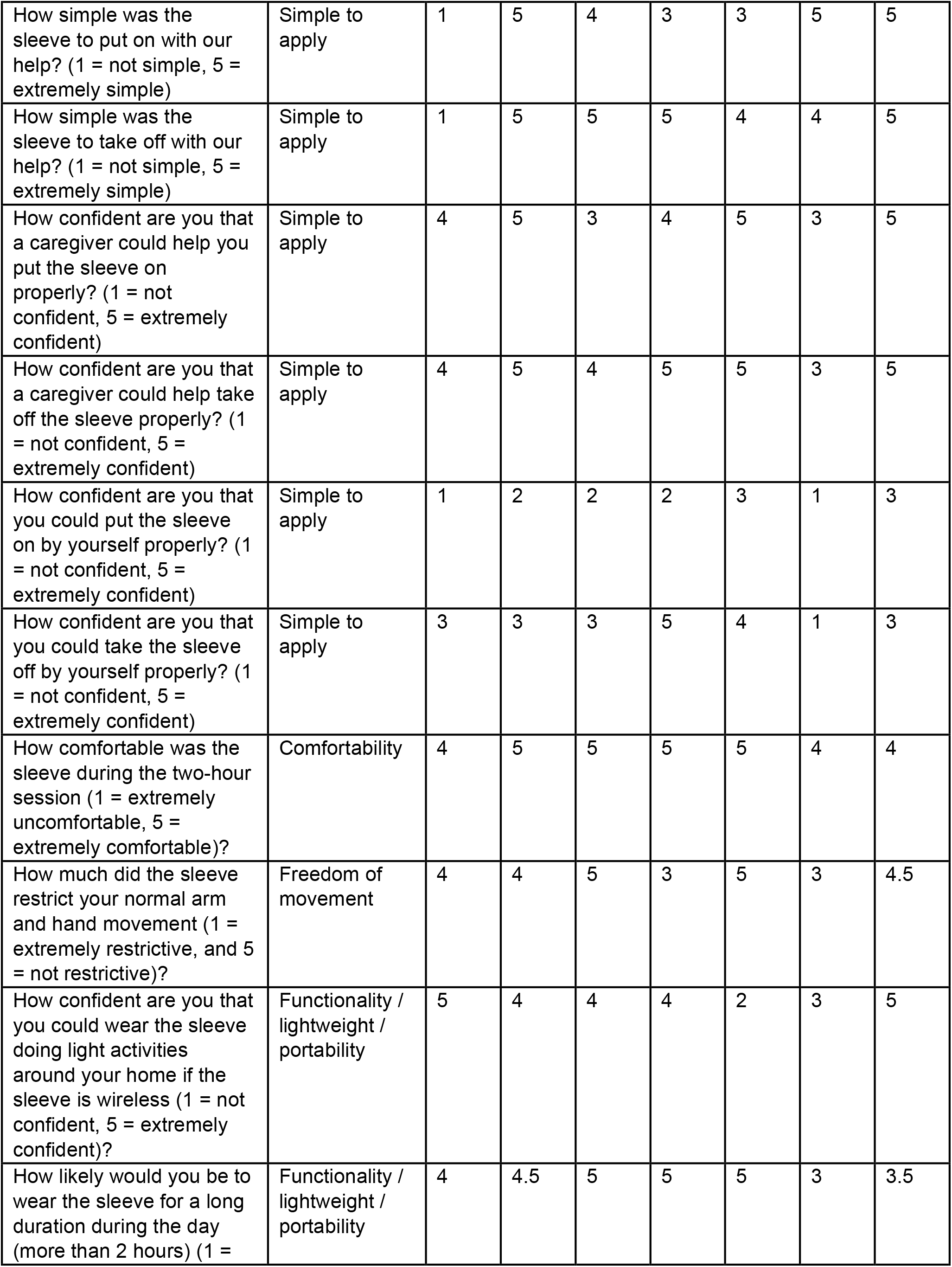
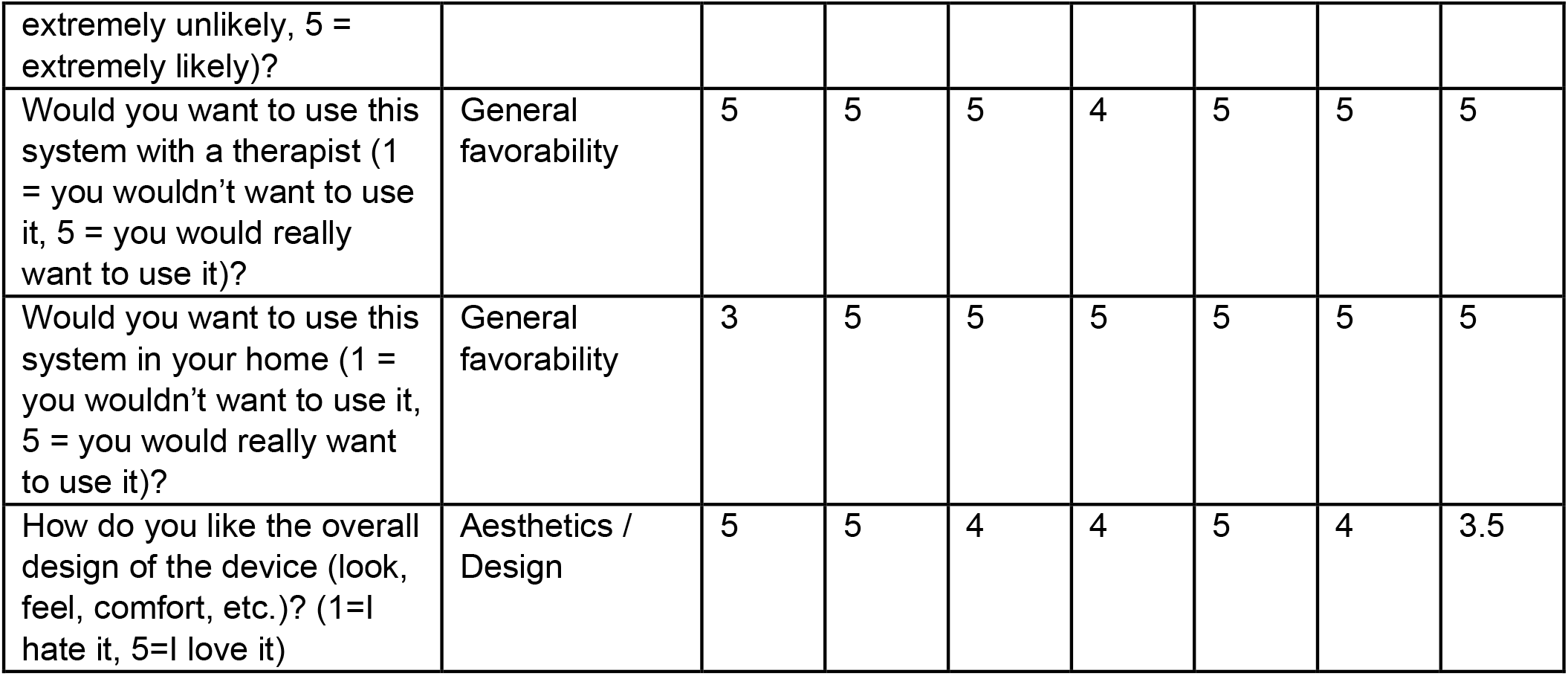
Complete usability questionnaire responses from all participants with stroke in the study. Each question was categorized to a specific domain, shown in the domain column and this data was used in Figure 6.

